# Benefits of hydrogen during constant load testing in healthy adults: A pilot double-blind randomized crossover trial

**DOI:** 10.1101/2025.10.26.25338854

**Authors:** Yuki Muramoto, Genta Toshima, Emi Minaguchi, Kazuhisa Sugai, Yuji Iwasawa, Kohsuke Shirakawa, Kengo Nagashima, Yasunori Sato, Motoaki Sano, Kazuki Sato, Yoshinori Katsumata

## Abstract

**Purpose:** Exercise-induced fatigue hinders activity. Molecular hydrogen may enhance mitochondrial function and efficiency, but its effects in non-athletes are unclear. We evaluated whether hydrogen-rich jelly (HRJ) enhances high-intensity exercise in non-athletes.

**Methods:** This double-blind, randomized, placebo-controlled crossover trial (UMIN000053818) enrolled 50 healthy adults, who were randomly assigned in a 1:1 ratio to ingest HRJ first or placebo jelly (PJ) first. Participants consumed 10 g of HRJ or PJ on three occasions: 24 h, 90 min, and immediately before exercise. An incremental test determined anaerobic threshold (AT), followed by 30 min of cycling at 120% of AT. Oxygen consumption (VO□) and heart rate (HR) were continuously monitored; blood lactate was sampled every 5 min. Condition differences were assessed with paired t-tests and mixed-effects models.

**Results:** Of the 50 participants, 48 completed all measurements, and analyses were restricted to the 28 who completed the entire 30-min exercise session (13 women, 46.4%). The mean age was 41.7 ± 11.5 years; mean BMI 22.3 ± 2.6 kg/m²; and mean peak VO□, 26.8 ± 5.9 mL/kg/min. In the PJ condition, maximum lactate reached 5.12 mmol/L. Compared with PJ, HRJ resulted in significantly lower lactate levels at 5 and 10 min, as well as a lower peak lactate (mean difference −0.75, p = 0.014). VO□ at 10 and 30 min and peak VO□ were also lower with HRJ (peak difference −1.17, p = 0.017). HR showed no significant difference between conditions.

**Conclusion:** HRJ reduced peak blood lactate and VO□, suggesting suppressed metabolite accumulation and improved exercise efficiency.

## 1. Introduction

A plethora of substantial benefits have been attributed to exercise, including weight reduction,^1,2^ improved exercise tolerance,^1,2^ and enhanced quality of life.^3,4^ Exercise plays a pivotal role in both the primary and secondary prevention of disease onset and progression, thereby reducing the overall mortality risk.^1–4^ To maximize these benefits, the World Health Organization (WHO) recommends approximately 75 min per week of vigorous-intensity exercise, defined as activity that results in perspiration and a sense of breathlessness.^5^ Importantly, vigorous-intensity exercise elicits greater improvements in oxygen consumption compared with low-intensity exercise^6^ and is consistently associated with a reduced risk of mortality.^7–9^ Thus, the health benefits of exercise have been demonstrated from multiple perspectives, and physical activity has been increasingly recommended for the general population in various guidelines and position statements.^1,4,5^ Consequently, there has been an observed trend of individuals increasingly participating in regular exercise as part of their daily routines.^10^ However, the recommended level of vigorous-intensity exercise corresponds to an intensity that partly relies on anaerobic metabolism for energy production and is often accompanied by fatigue, which limits the ability to sustain the prescribed workload.^11^ To facilitate the incorporation of relatively high-intensity exercise into daily life, strategies that enhance mitochondrial function (oxidative phosphorylation) and alleviate exercise-induced fatigue may be beneficial. Based on this rationale, we focused on molecular hydrogen.

Molecular hydrogen attenuates oxidative stress at the cellular level.^12^ Its efficacy and safety have been documented in both disease treatment and general health maintenance.^13,14^ Hydrogen-rich water (HRW), in which molecular hydrogen is dissolved, has been shown to reduce post-exercise lactate accumulation and alleviate muscle fatigue, with particularly notable effects in endurance athletes such as cyclists and marathon runners.^15–18^ One proposed mechanism involves the selective elimination of potent reactive oxygen species, such as hydroxyl radicals (•OH), via heme proteins.^19^ In addition, molecular hydrogen enhances mitochondrial membrane potential, thereby promoting mitochondrial activity and ATP production.^20^ Collectively, these processes are thought to suppress excessive lactate production. Physiological responses to high-intensity exercise have been extensively examined in athletes. However, much of the existing evidence derives from athlete cohorts, and the physiological effects of molecular hydrogen during exercise in non-athletes remain insufficiently explored. Recently, the methods of hydrogen administration have diversified. Our laboratory developed a technique to encapsulate molecular hydrogen within a jelly matrix composed of polysaccharides and gelatin, achieving a higher hydrogen concentration than conventional HRW.^21^ Each 10 g portion of this high-concentration hydrogen-rich jelly (HRJ) delivers hydrogen equivalent to approximately 600 mL of HRW. HRJ therefore represents a novel approach to daily hydrogen intake, offering greater convenience and efficiency than traditional HRW.

In this study, we investigated the effects of HRJ on metabolic responses during high-intensity exercise in non-athletes with moderate fitness levels. We hypothesized that HRJ intake would reduce inefficient oxygen utilization, suppress exercise-induced lactate accumulation, and thereby promote more efficient metabolic control from the perspective of energy production.

## 2. Materials and Methods

### 2.1. Hydrogen gas content in the hydrogen jelly

The HRJ used in this study was produced in a facility certified under the FSSC 22000 standard, a globally recognized food safety certification scheme approved by the Global Food Safety Initiative. The jelly contained uniformly dispersed molecular hydrogen, and each portion was individually packaged in aluminum pouches to preserve hydrogen content. The concentration of dissolved hydrogen gas was stable for up to 8 months (Supplemental Fig. 1, Supplemental Digital Content, Hydrogen molecule content in hydrogen-rich jelly). Using the headspace gas chromatography method, we confirmed that hydrogen levels remained above 30 ppm for up to 18 months after production.

The HRJ was stored at an ambient temperature of 25 °C, a safe and stable condition that preserved the integrity of the hydrogen concentration in the product.

### 2.2. Sample size

This study represents a secondary analysis focusing on exercise-related outcomes of HRJ. The sample size was determined for the primary analysis, which evaluated heart rate variability (HRV), with emphasis on the high-frequency (HF) component as an indicator of parasympathetic nervous system activity.^22^ Based on preliminary data, the required sample size to detect significant changes in HF was calculated using G*Power with a two-tailed α of 0.05 and a power (1–β) of 0.90, resulting in 36 participants. However, as approximately 20% of participants in the pilot data exhibited HF values exceeding three standard deviations above the mean (indicating potential outliers), the target sample size was increased to 45. Accounting for an anticipated dropout rate of 10%, the final recruitment target was set at 50 participants. For this secondary analysis, the dataset included 28 participants who successfully completed the exercise test from within this predefined sample.

### 2.3. Participants

Fifty healthy adults with varying fitness levels, independent of their exercise habits, were recruited between May and November 2024. Participants were aged 22 to 60 years, representing a broad demographic range. Inclusion criteria were as follows: absence of cardiovascular, respiratory, or metabolic diseases; absence of athletic injuries; non-smoking status; and no use of dietary supplements or medications for at least 1 week before participation. The study protocol was approved by the Institutional Review Board of Keio University School of Medicine (approval number: 20231194) and conducted in accordance with the principles of the Declaration of Helsinki. All participants were informed about the study and provided written informed consent. The study was registered in the University Hospital Medical Information Network (UMIN000053818).

### 2.4. Study design

This study employed a double-blind, randomized, placebo-controlled crossover design. The study protocol consisted of three sessions **(Visit 1, 2, and 3; Fig. 1)**. On the first day (Visit 1), body weight, body fat, and skeletal muscle mass were assessed using a bioelectrical impedance analyzer (Inbody 470; Inbody Japan Inc., Tokyo, Japan). The anaerobic threshold (AT) was then determined with a ramp load test for exhaustion, performed on an electromagnetically braked ergometer (StrengthErgo8 V2; Fukuda Denshi Co., Ltd., Tokyo, Japan) equipped with an exhaled gas analyzer (Aeromonitor®, MINATO Medical Science Co., Ltd., Osaka, Japan) and a heart rate (HR) monitor (Polar H10, Polar Electro Japan Co., Ltd., Tokyo, Japan). After AT determination, participants returned on two separate occasions to consume either 10 g of HRJ (30–40 ppm; Shinryo Co., Ltd., Fukuoka, Japan) or placebo jelly (PJ), followed by a 30-min constant-load test at 120% of AT **(Visits 2 and 3; Fig. 1).** In addition, exhaled acetone (Ex-ace) was collected from 20 consenting participants before and after exercise using the same device (ReCIVA Breath Sampler).

**Fig. 1.**
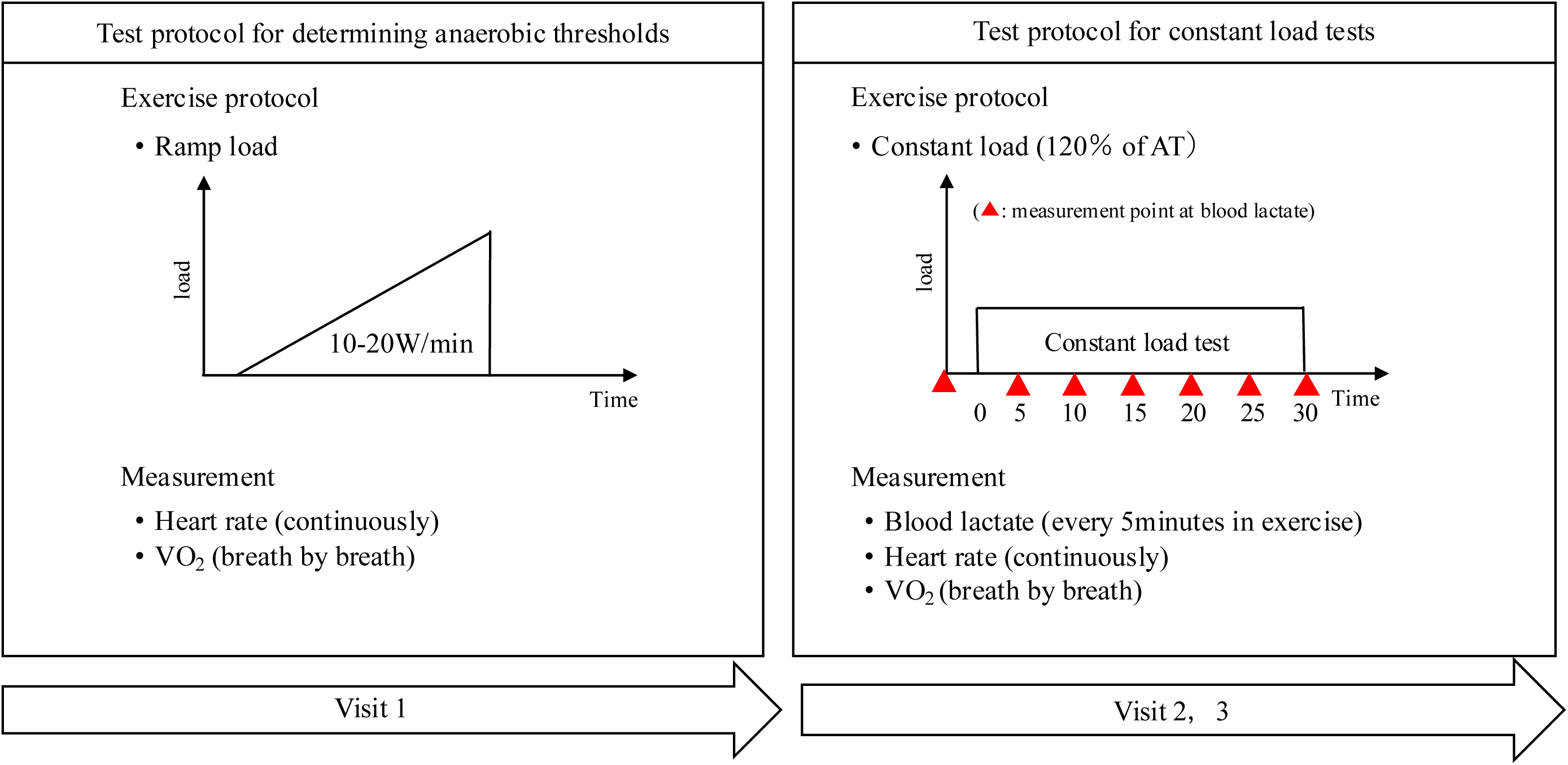
Protocols for tests 1, 2, and 3. Red triangle: measurement point of blood lactate; W·min^-1^: workload per min; VO□ : oxygen uptake. This study was conducted as a double-blind, randomized, placebo-controlled crossover trial consisting of three sessions (Visits 1–3). At Visit 1, body composition (body weight, body fat, and skeletal muscle mass) was measured using bioelectrical impedance (Inbody 470; Inbody Japan Inc., Tokyo, Japan), and the anaerobic threshold (AT) was determined using a ramp load test to exhaustion (StrengthErgo8 V2; Fukuda Denshi Co., Ltd., Tokyo, Japan) with an exhaled gas analyzer (Aeromonitor®, MINATO Medical Science Co., Ltd., Osaka, Japan) and a heart rate (HR) monitor (Polar H10, Polar Electro Japan Co., Ltd., Tokyo, Japan). At Visits 2 and 3, participants consumed either 10 g of HRJ (30–40 ppm; Shinryo Co., Ltd., Fukuoka, Japan) or placebo jelly (PJ) in random order, followed by a 30-min constant-load exercise test at 120% AT.

Participants were randomly assigned (1:1) to two groups using an allocation sequence generated in Microsoft Excel, which determined whether HRJ or PJ was administered first. Administration followed a double-blind design, with containers labeled ID_1 or ID_2. Participants consumed the ID_1 drink before Visit 2 and the ID_2 drink before Visit 3, ensuring that neither the participants nor the investigators could identify the hydrogen water condition. The two conditions were measured with a washout interval of at least 7 days. The concentration of hydrogen in the body reaches its peak within a few minutes after hydrogen gas inhalation,^16^ and hydrogen in the exhaled breath is completely lost 60 min after intake.^17^ Therefore, participants were instructed to drink 10 g of HRJ or PJ within 2 min at the following time points: 24 h, 90 min, and immediately before the test.

### 2.6. Ramp test protocol

Participants were asked to fast for 3 h before the test and were prohibited from consuming caffeine, which can enhance exercise performance by mitigating or delaying fatigue, and alcohol, which may accelerate fatigue, for 12 h before the test. After measuring the remaining baseline data for 2 min, the participants performed a warm-up exercise for 2 min at a 20-W load and then exercised at increasing intensities until they could no longer maintain the pedaling rate (volitional exhaustion). The load was increased by 15 W at 1-min intervals. The rotational speed was maintained at 70 rpm.

### 2.7. Respiratory gas analysis and AT

Expired gas flow was assessed with a breath-by-breath automated system, and respiratory gas exchange variables (minute ventilation [VE], oxygen uptake [VO_2]_, and carbon dioxide output [VCO_2_]) were continuously monitored and averaged over 10 s. A three-step calibration process was performed for the flow-volume sensor, including delay-time adjustment. Peak VO_2_ was measured as the average oxygen consumption during the last 30 s of exercise. The ventilatory efficiency slope (VE-VCO_2_ slope) was calculated using linear regression of data collected from the start of exercise until the respiratory compensation point. Initial HR was defined as the average heart rate recorded during the 2 min preceding exercise in a seated position. AT was assessed using the ventilatory equivalent, excess carbon dioxide, and modified V-slope methods.^23^ AT decisions were made by experienced cardiac rehabilitation doctors and physical therapists.

### 2.8. Constant load test protocol

This test was conducted at 120% of the AT workload for 30 min. The rotational speed was set at 70 rpm. During the constant-load exercise, an exhaled gas analyzer and an HR monitor were attached. Blood lactate values were obtained by auricular pricking and gentle squeezing of the earlobe using a blood lactate analyzer (Lactate Pro 2, ARKRAY Inc., Kyoto, Japan). Blood lactate concentrations were measured at baseline and at 5-min intervals during the 30-min exercise bout. Exercise termination was defined as the inability to sustain pedaling at 70 rpm because of fatigue.

### 2.9. Exhaled Acetone Sampling and Acetone Quantification

Exhaled acetone (Ex-ace) was collected according to a standardized protocol using the Breath Biopsy® platform (Owlstone Medical). Sampling was performed with the ReCIVA® Breath Sampler, coupled with the CASPER® Portable Air Supply, and breath was adsorbed onto Tenax TA/Carbograph 5TD-packed tubes (Markes International, Llantrisant, UK). Breathing patterns were monitored in real time using CO□ concentration and pressure sensors, and breath sampling was triggered when the exhaled CO□ concentration reached approximately 0.6%. Approximately 1.0 L of exhaled breath was collected over 8–12 min at a flow rate of 225 mL·min^-1^, adjusted to each participant’s tidal volume. Ambient contamination was minimized by using the CASPER® Portable Air Supply. Before sampling, the adsorbent tubes were conditioned at 280 °C under a nitrogen flow (20 psi) for 30 min. After collection, breath samples were stored at −40 °C and analyzed the next day using a Shimadzu TD-30R thermal desorption system (Shimadzu Corporation, Kyoto, Japan) coupled to a GCMS-QP2010 Ultra mass spectrometer. For Ex-ace peak analysis, individual peaks were identified using a pre-established system library based on retention indices and mass spectra. Optimized integration parameters were applied to each peak, and the corresponding peak areas were calculated.

### 2.10. Statistical analysis

Continuous variables were summarized as means with standard deviations, while categorical variables were expressed as frequencies with percentages.

Differences in HR, VO_2_, and blood lactate levels between conditions during constant-load exercise were analyzed using mixed-effects models for repeated measures. These differences in HR, VO□, and lactate changes were assessed using paired t-tests and mixed-effects models for repeated measures, with 95% confidence intervals (CIs) estimated.

For VOC analysis, among the 20 participants whose VOCs were successfully measured, nine participants who completed the constant-load exercise were included for comparison. Exhaled acetone peak area changes were compared between HRJ and PJ conditions using paired t-tests and mixed-effects models, consistent with the analyses of HR, VO□, and lactate.

Additional subgroup analyses were performed to explore participant characteristics associated with the difference in HR, VO_2,_ and blood lactate levels changes between the HRJ and PJ conditions. Subgroups were defined according to nine characteristics: sex (male/female), smoking status (no/yes), alcohol intake (daily alcohol consumption of < 20 g / ≥ 20 g), physical activity habits (<3 times per week / ≥3 times per week and <4 times per week / ≥4 times per week), age (< 47 / ≥ 47), body mass index (BMI) (<22.8 kg·m^-^² / ≥22.8 kg·m^-^²), body fat percentage (< 24.9% / ≥ 24.9%), and skeletal muscle mass (<40 kg / ≥40 kg). Cutoff values for BMI, body fat percentage, skeletal muscle mass, and age were determined from baseline sample medians.

Effect sizes were calculated using Cohen’s d, based on within-subject mean differences and standard deviations. All p-values were two-tailed, and statistical significance was set at p < 0.05. Statistical analyses were performed using SAS software, version 9.4 (SAS Institute, Japan).

## 3. Results

Fifty participants were initially recruited for the study; however, two withdrew prior to commencement due to scheduling conflicts. Among the remaining participants, 48 (27 females) completed the HRJ and PJ interventions and participated in both the constant-load exercise test (Table 1). However, 20 participants were unable to complete the constant-load exercise for 30 min and were excluded from the primary statistical analysis. Thus, the analysis included data from 28 participants (Figure 2).

**Fig. 2.**
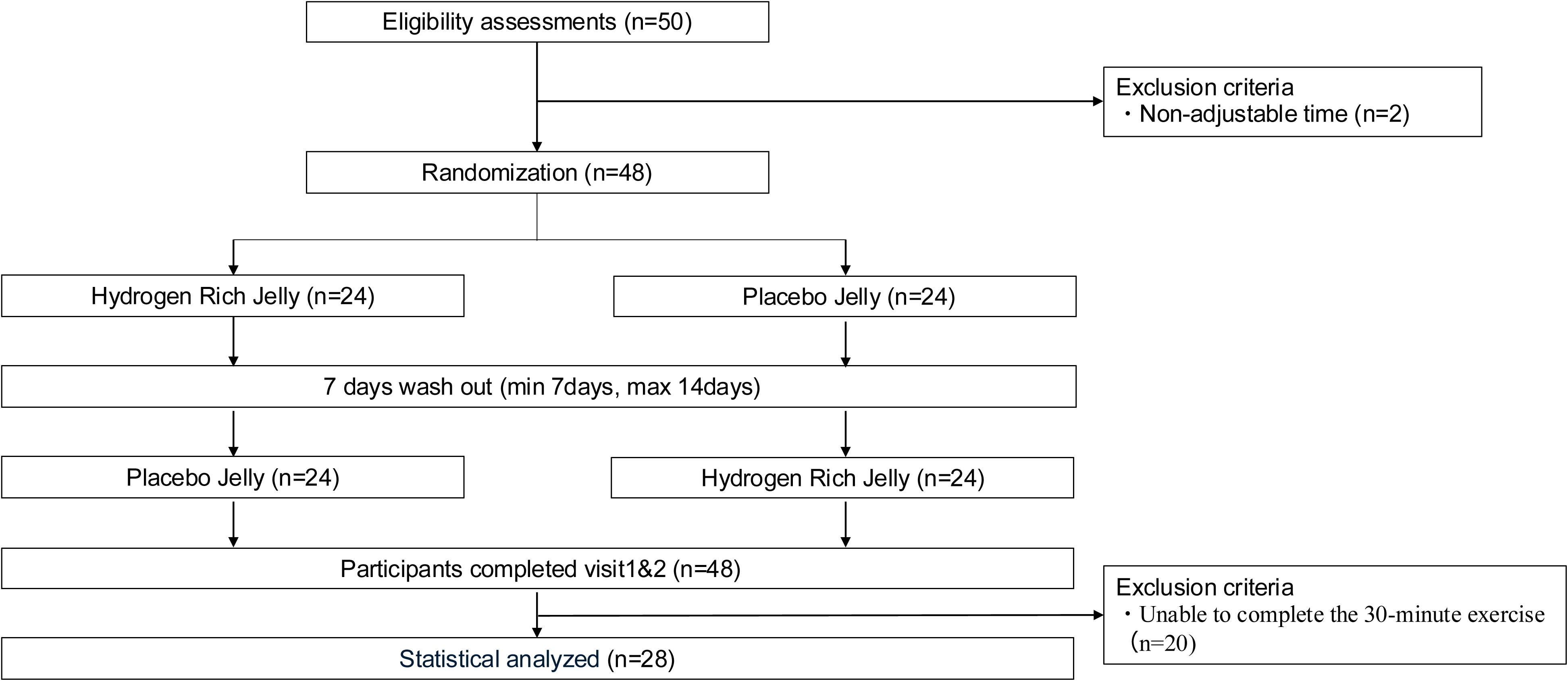
CONSORT flow diagram of the study protocol. Fifty participants were initially recruited, of whom two withdrew before the start of the study due to scheduling conflicts. The remaining 48 participants completed both the hydrogen-rich jelly (HRJ) and placebo jelly (PJ) interventions and underwent the constant-load exercise test. Among them, 20 participants were unable to complete the 30-min constant-load exercise and were excluded from the primary statistical analysis. Consequently, data from 28 participants were included in the final analysis.

### 3.1. Effects of HRJ on Metabolic Responses During Exercise

HRJ intake significantly reduced blood lactate levels and VO□ during exercise compared with PJ. Blood lactate concentrations were notably lower under the HRJ condition at 5 min (mean difference = –0.49, p = 0.004) and 10 min (mean difference = –0.63, p = 0.008). A trend toward lower values was also observed at 25 min (mean difference = –0.48, p = 0.076). (Table 2, Fig.3). Additionally, peak blood lactate levels during exercise were significantly lower in the HRJ condition (mean difference = –0.75, p = 0.014; Table 2, Fig.3). Similarly, VO□ was significantly lower in the HRJ condition at 10 min (mean difference = –1.20, p = 0.016) and 30 min (mean difference = –1.40, p = 0.017). A trend toward lower VO□ was also observed at 25 min (mean difference = –1.13, p = 0.081; Table 2, Fig. 4). Additionally, peak VO□ during constant-load exercise was also significantly lower with HRJ than with PJ (mean difference = –1.17, p = 0.014; Table 2, Fig. 4).No significant differences were observed between the two conditions in HR at any time point or in peak HR during exercise (Table 2, Fig.5). Regarding the peak area of Ex-ace that emerged before and after exercise, a trend toward a lower response was observed with HRJ than with PJ (mean difference = –4,129,905, p = 0.090).

**Fig. 3.**
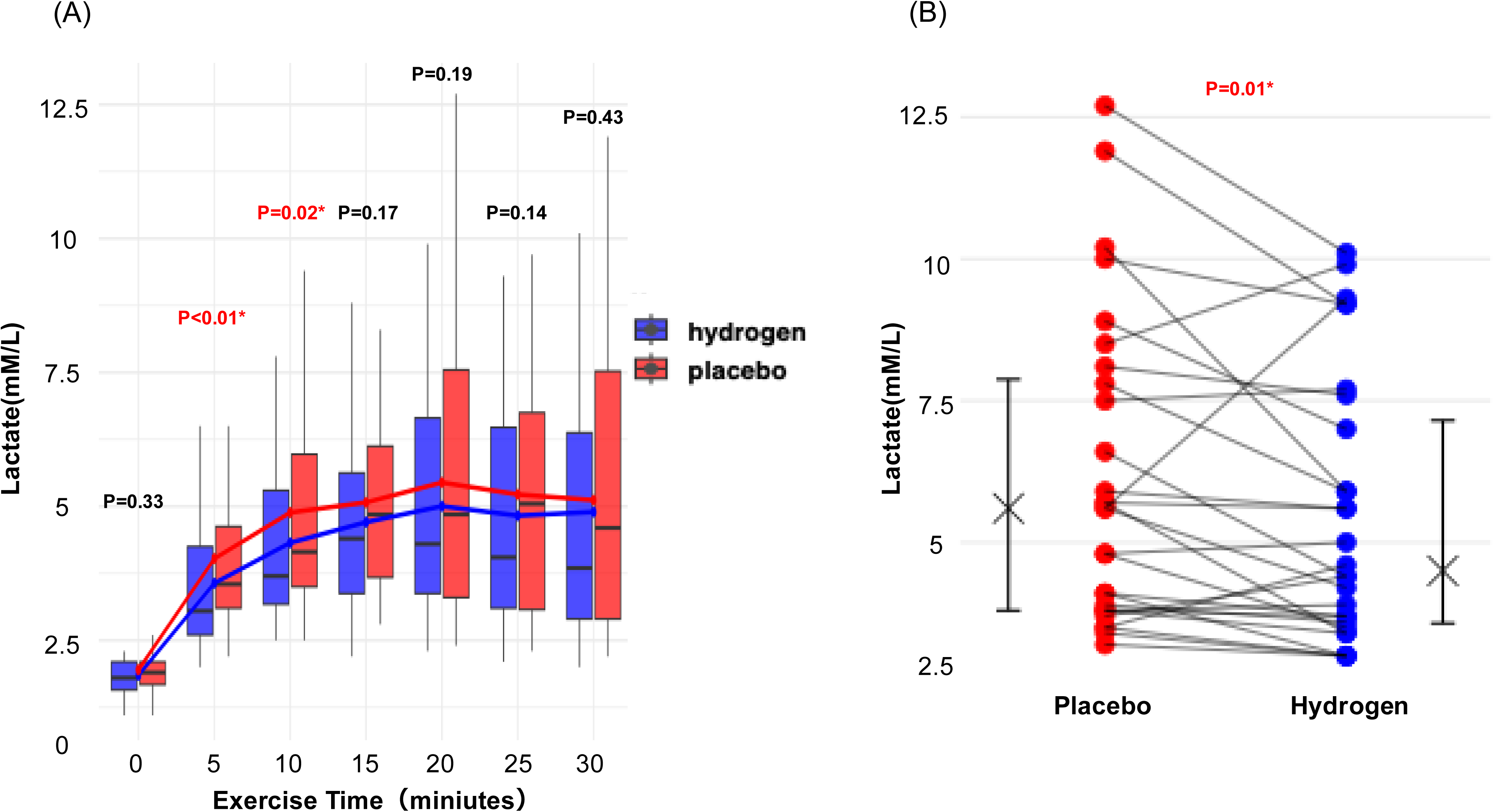
Blood lactate responses during the constant-load exercise test. (A) Time course of blood lactate concentrations measured every 5 min during exercise under the hydrogen-rich jelly (HRJ) and placebo jelly (PJ) conditions. Blood lactate concentrations were significantly lower with HRJ at 5 min (mean difference = –0.49, p = 0.004) and 10 min (mean difference = –0.63, p = 0.008), with a trend toward lower values at 25 min (mean difference = – 0.48, p = 0.076). (B) Peak blood lactate levels during exercise. Peak values were significantly lower with HRJ than with PJ (mean difference = –0.75, p = 0.014). Data are presented as mean ± SD. *p < 0.05 vs. PJ.

**Fig. 4.**
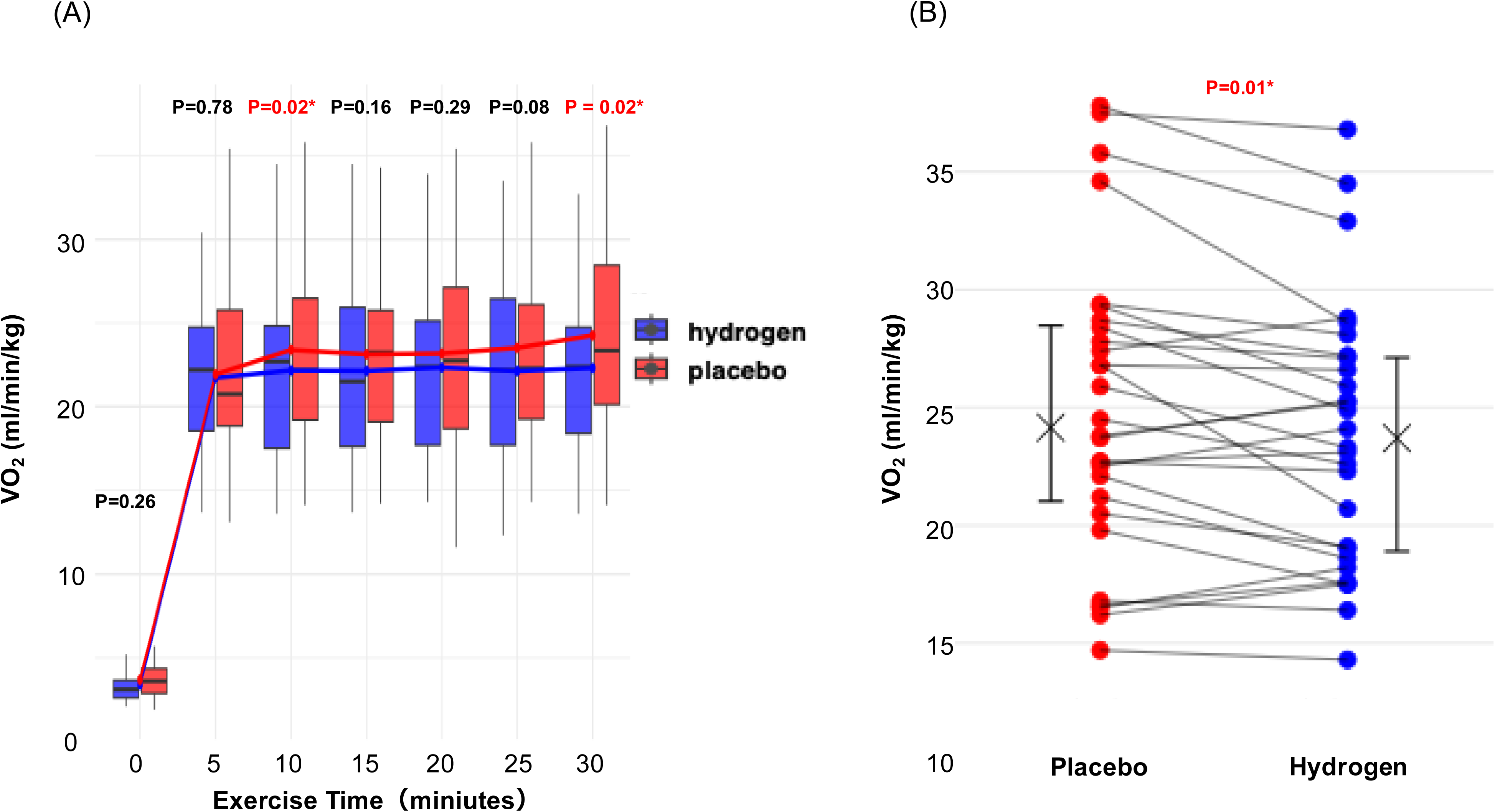
Oxygen consumption during the constant-load exercise test. (A) Time course of oxygen uptake (VO□) measured every 5 min during exercise under the hydrogen-rich jelly (HRJ) and placebo jelly (PJ) conditions. VO□ was significantly lower in the HRJ condition at 10 min (mean difference = –1.20, p = 0.016) and 30 min (mean difference = – 1.40, p = 0.017), with a trend toward lower values at 25 min (mean difference = –1.13, p = 0.081). (B) Peak VO□ during exercise. Peak values were significantly lower with HRJ than with PJ (mean difference = –1.17, p = 0.014). Data are presented as mean ± SD. *p < 0.05 vs. PJ.

**Fig. 5.**
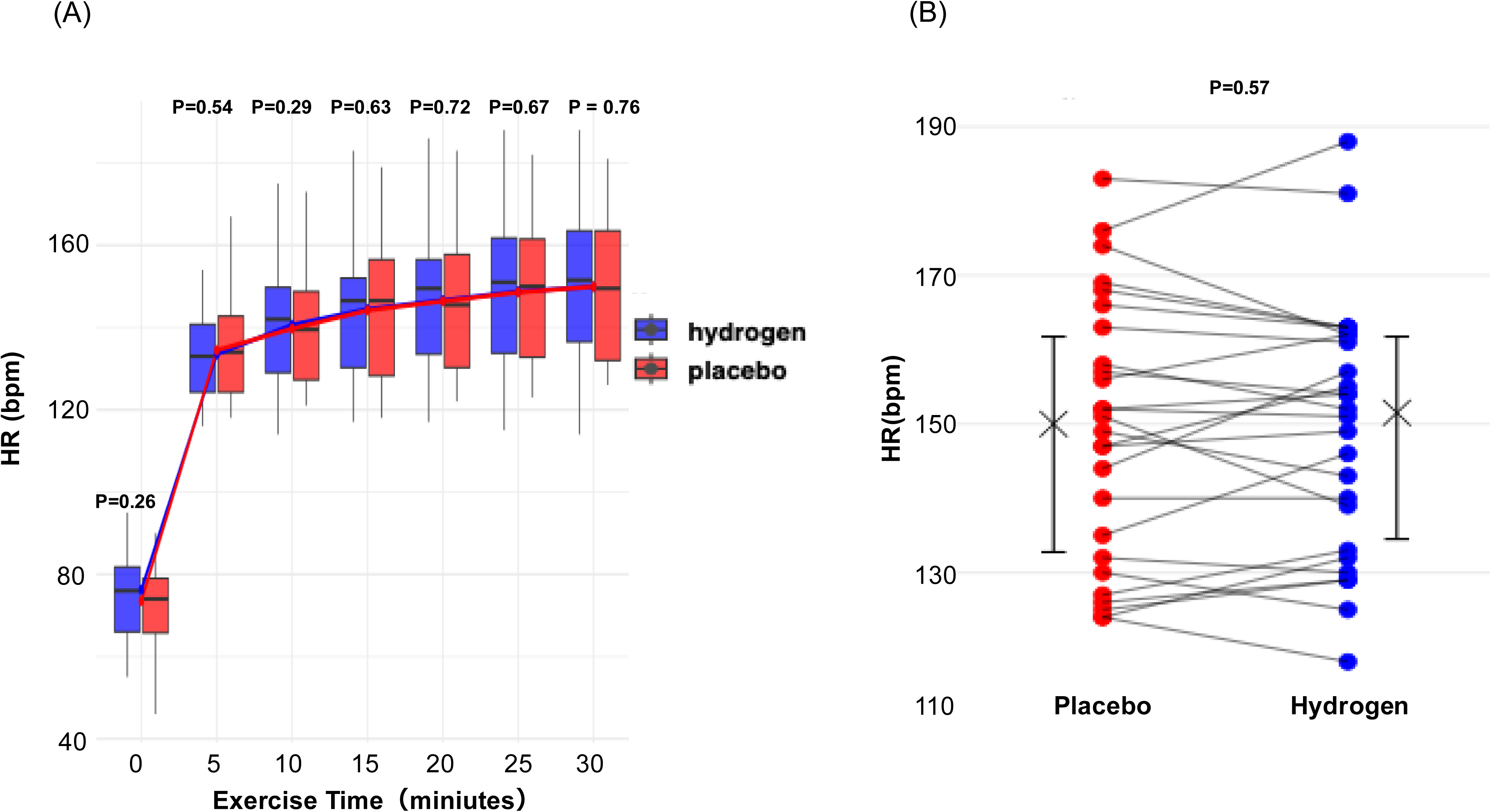
Heart rate during the constant-load exercise test. (A) Time course of heart rate (HR) measured every 5 min during exercise under the hydrogen-rich jelly (HRJ) and placebo jelly (PJ) conditions. (B) Peak HR during exercise. No significant differences were observed between the two conditions at any time point or in peak HR. Data are presented as mean ± SD.

### 3.2. Participant Characteristics Associated with the Impact of HRJ on Peak Exercise Response

Participants who exhibited a reduction in peak blood lactate levels under the HRJ condition were primarily characterized by being female (mean difference = –0.90, 95% confidence interval [CI]: –1.64 to –0.15, p = 0.023), daily alcohol consumption ≥20 g (mean difference = –0.86, 95% CI: –1.53 to –0.20, p = 0.014), exercising less than three times per week (mean difference = – 0.75, 95% CI: –1.27 to –0.23, p = 0.008), and body fat percentage of ≥24.9% (mean difference = –1.09, 95% CI: –1.93 to –0.25, p = 0.018). Participants younger than 47 years also showed a trend toward a greater reduction (mean difference = –0.84, 95% CI: –1.72 to 0.03, p = 0.058), although this did not reach statistical significance (Supplemental Fig. 2, Supplemental Digital Content, Characteristics of participants who exhibited a reduction in peak blood lactate under the hydrogen-rich jelly (HRJ) condition).

Participants showing a reduction in peak VO□ under the HRJ condition were characterized by a BMI of <22.8 kg·m^-^² (mean difference = –1.75, 95% CI: –2.83 to –0.68, p = 0.004), body fat percentage <24.9% (mean difference = –1.58, 95% CI: –2.70 to –0.47, p = 0.009), skeletal muscle mass ≥40 kg (mean difference = –1.45, 95% CI: –2.81 to –0.10, p = 0.037), and a habitual exercise frequency ≥4 times per week (mean difference = –1.51, 95% CI: –2.79 to – 0.22, p = 0.033). Male participants (mean difference = –1.59, 95% CI: –3.24 to 0.07, p = 0.059) and participants aged ≥47 years (mean difference = –1.39, 95% CI: –2.96 to 0.18, p = 0.075) also showed a trend toward greater reductions, although these differences did not reach statistical significance (Supplemental Fig. 3, Supplemental Digital Content, Characteristics of participants who exhibited a reduction in peak oxygen uptake (VO□) under the hydrogen-rich jelly (HRJ) condition.).

## 4. Discussion

This study examined the effects of HRJ intake on physiological responses to high-intensity exercise in non-athletes. Our findings corroborate those of earlier studies, showing that HRJ blunted the rise in blood lactate during the first 10 min of exercise and reduced peak lactate levels during exercise. A novel observation was that peak VO□ during exercise was significantly lower with HRJ. Collectively, these findings suggest that HRJ reduces unnecessary oxygen utilization and attenuates lactate accumulation, thereby improving exercise efficiency in non-athletes, even at comparable intensities (Fig.6).

**Fig. 6.**
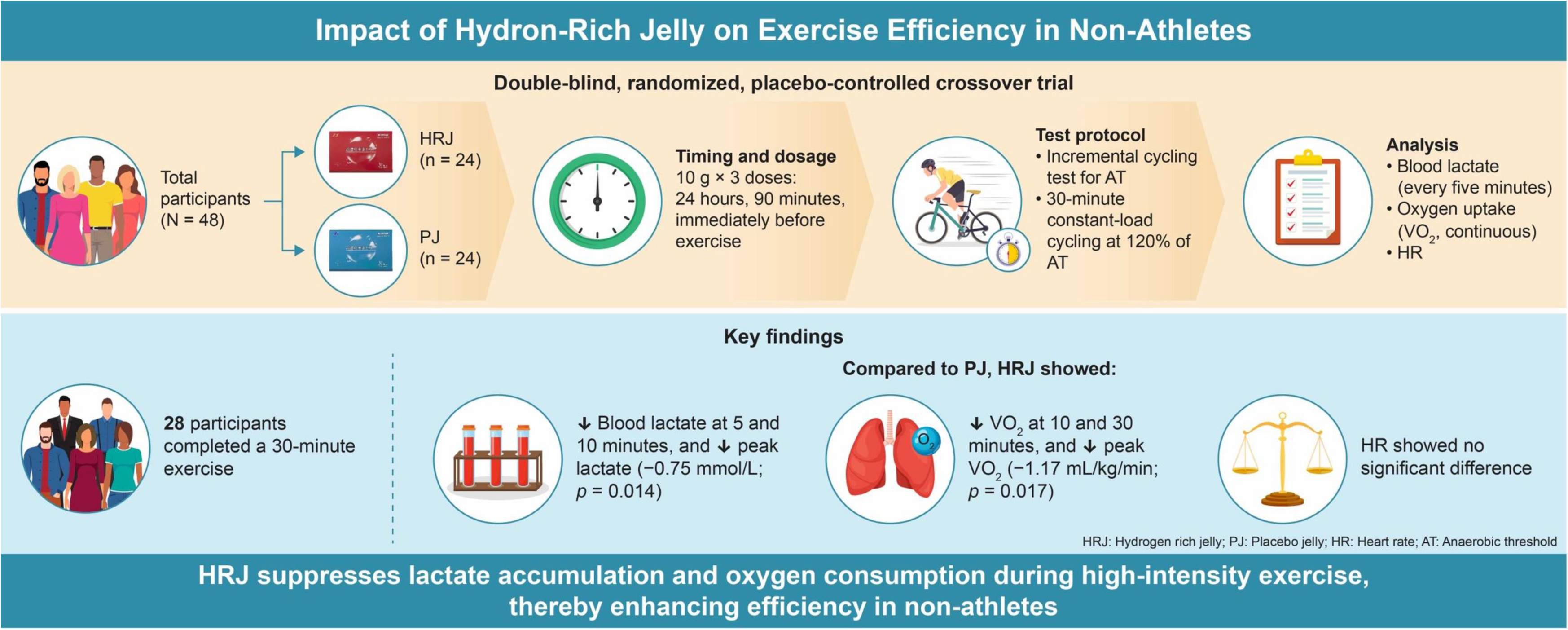
Graphical summary of the effects of hydrogen-rich jelly (HRJ) intake on exercise efficiency during high-intensity exercise in non-athletes. A double-blind, randomized, placebo-controlled crossover trial was conducted with 48 participants, and data from 28 participants who completed the 30-min constant-load exercise were analyzed. Participants ingested HRJ or placebo jelly (PJ, 10 g × 3 doses: 24 h, 90 min, and immediately before exercise), performed an incremental cycling test to determine the anaerobic threshold (AT), and then completed 30 min of constant-load cycling at 120% of AT. Compared with PJ, HRJ suppressed the rise in blood lactate at 5 and 10 min and significantly reduced peak lactate levels (–0.75 mmol·L^-1^, *p* = 0.014). In addition, VO□ was significantly lower at 10 and 30 min as well as at peak values (–1.17 mL·min^-1^·kg^-1^, *p* = 0.017). No significant difference was observed in heart rate between conditions.

### 4.1. Effects of HRJ on Metabolic Responses During Exercise

Previous studies have reported that HRW suppressed blood lactate accumulation in athletes during high-intensity exercise, such as that performed at 75% of peak VO□ or during the Quebec 90-s test.^15,16^ In the current trial, the mean blood lactate concentration under the placebo condition was 5.12 mmol·L^-1^, which was within the range of 5–12 mmol·L^-1^ observed in previous studies.^15,16^ These results indicated that, even in non-athletes, aerobic exercise of sufficient intensity to elevate lactate levels to a comparable range revealed the lactate-suppressing effect of hydrogen gas. Here, the lactate level increase within the first 10 min of exercise and the peak lactate level were attenuated. At the onset of high-intensity exercise, oxygen supply falls short of demand, resulting in an oxygen deficit. Consequently, glycogen is anaerobically broken down to generate ATP, leading to lactate accumulation.^23,24^ Lactate is subsequently reutilized as a substrate for ATP resynthesis within mitochondria, thereby supporting sustained exercise performance.^25^ In addition, lactate produced within skeletal muscles is transported to the liver and converted to glucose via the Cori cycle, which is then reutilized as an energy substrate.^26^ Thus, suppression of lactate elevation during high-intensity exercise could be attributed to both reduced lactate production and enhanced lactate utilization in skeletal muscles, as well as augmented gluconeogenic capacity in hepatocytes. With respect to the former, the effect was likely mediated by improved mitochondrial function within skeletal muscles. Our findings also showed that HRJ intake suppressed VO□ during exercise. This suggests that mitochondrial oxygen utilization efficiency was enhanced, thereby allowing the same exercise intensity to be maintained with lower oxygen consumption. Skeletal muscle mitochondria generate energy through oxygen consumption; however, during this process, superoxide (O□□) and hydrogen peroxide (H□O□) are produced. In the presence of transition metals, such as iron (Fe), the Fenton reaction generates highly reactive hydroxyl radicals (•OH), which subsequently trigger lipid peroxidation.^27^ More so, lipid peroxide accumulation impairs mitochondrial oxygen uptake capacity.^28^ Molecular hydrogen selectively neutralizes hydroxyl radicals (•OH) and suppresses lipid peroxidation chain reactions.^12,19,29^ This antioxidative action prevents a decline in oxygen utilization efficiency, thereby contributing to the observed reduction in VO□. Such improvements in mitochondrial function were also likely to reduce skeletal muscle lactate production and facilitate energy production via lactate reutilization. Generally, hydrogen gas inhaled via the gastrointestinal tract is absorbed and almost completely exhaled through the lungs.^30^ However, during exercise, the shortened pulmonary capillary transit time and increased intrapulmonary shunting reduced hydrogen exhalation, suggesting that hydrogen gas might have remained in the arterial circulation. Future studies are required to investigate the hemodynamics of hydrogen gas during exercise in greater detail.

HRJ intake also tended to suppress the post-exercise rise in Ex-ace. High-intensity exercise typically results in depletion of muscle glycogen stores, leading to a metabolic shift toward fatty acid oxidation.^31,32^ Consequently, ketone bodies are generated in the liver, with acetone being produced as a byproduct.^31,32^ Therefore, the attenuated increase in Ex-ace under the HRJ condition suggests that improved oxygen utilization efficiency promoted glucose metabolism and lactate reutilization while limiting excessive reliance on fatty acid oxidation. Orally ingested molecular hydrogen delivers high concentrations of hydrogen to the liver via the portal vein.^30,33,34^ The liver plays a key role in lactate reutilization through the Cori cycle, and improvement of hepatic function induced by hydrogen intake likely contributed to the enhanced lactate clearance. Furthermore, in the present study, HRJ was ingested 24 h before exercise, as well as 90 min and immediately before exercise. Considering hydrogen gas rapidly disappears from the body after ingestion, acute effects of hydrogen gas on the organs were unlikely present 24 h after ingestion. Instead, a preconditioning effect might have been induced via activation of the PGC-1α pathway, promoting functional improvement of existing mitochondria and stimulating mitochondrial biogenesis.^33^ Thus, both skeletal muscle and liver metabolism were primed, resulting in more pronounced suppression of lactate accumulation and improvement in oxygen utilization efficiency.

### 4.2. Participant Characteristics Associated with the Impact of HRJ on Peak Exercise Response

We examined the characteristics of participants whose peak blood lactate concentration and peak VO□ were reduced by HRJ intake during exercise. The reduction in peak lactate was significant primarily in women, individuals exercising less than three times per week, those with body fat percentages above the median, and those consuming more than 20 g of alcohol per day. Conversely, peak VO□ reduction was significant mainly in men with greater skeletal muscle mass (≥40 kg), below-median BMI and body fat percentages, and regular exercise four or more times per week.

A plausible explanation for these sex- and body composition-related differences is variations in skeletal muscle fiber composition and metabolic properties. Generally, women possess a higher proportion of type I (slow-twitch) fibers, characterized by superior oxidative metabolic capacity, whereas men typically have a higher proportion of type II (fast-twitch) fibers, associated with greater reliance on anaerobic metabolism and higher oxygen consumption during high-intensity exercise.^35,36^ Beyond fiber distribution, sex differences have also been reported in lactate production and clearance, with women exhibiting a greater capacity for lactate reutilization and more efficient lactate removal during recovery compared with men.^37^ Molecular hydrogen has been shown to enhance mitochondrial function.^20^ Accordingly, it is plausible that lactate production during exercise was suppressed in women through mitochondrial action, thereby reducing metabolic load. In addition, individuals with higher body fat percentages or habitual alcohol consumption are prone to elevated oxidative stress; however, HRJ intake may have strengthened antioxidant defense mechanisms, which in turn could have contributed to the suppression of lactate elevation.

Conversely, reductions in peak VO□ were primarily observed in men with greater skeletal muscle mass and more frequent exercise habits. These individuals are generally predisposed to higher oxygen consumption during muscle activity; however, HRJ intake may have improved the efficiency of oxygen delivery and utilization, thereby reducing the VO□ required to sustain the same exercise intensity. Molecular hydrogen has also been reported to improve blood flow,^38^ suggesting that enhanced peripheral oxygen delivery through vasodilation and improved microcirculation may have contributed to this outcome. These circulatory improvements could further enhance muscular oxygen utilization, ultimately reducing the VO□ required to sustain exercise performance.

### 4.3. Limitation

This study has several limitations. First, it was designed to examine whether HRJ influences physiological indices during high-intensity exercise in non-athletes. As a result, nearly half of the participants did not complete the study protocol. It remains unclear whether HRJ provides beneficial effects during moderate, daily life exercise in non-athletes. Second, the present study investigated only the acute effects of HRJ intake, and its long-term impact on exercise performance and physiology is still unknown. Further studies are warranted to elucidate the potential long-term physiological and performance-related effects of HRJ. Third, the analysis of Ex-ace was based on a small analyzable sample (n = 9), limiting statistical power. A power analysis performed with G*Power 3.1 estimated that 16 participants would be necessary to achieve 80% statistical power at a two-sided α of 0.05, highlighting the potential risk of type II error with the current sample. Exercise performance is also influenced by daily physiological variations, which could contribute to inter-session variability.

Future studies should address these limitations by examining the effects of HRJ during exercise at daily life intensities, evaluating the impact of longer-term intake, and incorporating analyses that consider participants’ fitness levels. Such approaches will help to more clearly elucidate the potential physiological effects of HRJ.

## 5. Conclusions

In the present study, intake of HRJ led to reductions in both peak blood lactate concentration and peak VO□. Furthermore, HRJ was associated with a suppression of the rise in Ex-ace, suggesting that excessive metabolite production may be attenuated even at the same exercise intensity. Collectively, these findings indicate that HRJ may enhance exercise efficiency and reduce metabolic load, providing a potentially useful strategy even for non-athletes.

## Acknowledgements

This work was supported by Shinryo CO., LTD. However, this corporation was not involved in data analysis and manuscript writing. The authors would like to thank the participants who actively participated in this study and SOUKEN Corporation (https://www.souken-lab.co.jp/) for their help with recruitment. The manuscript has been edited by Editage (www.editage.jp). The authors express their sincere appreciation to Noriko Akiyama for her substantial contributions to the measurement of VOC.

## Ethical Statement

The study protocol was approved by the Institutional Review Board of the Keio University School of Medicine (approval number: 20221205) and was conducted in accordance with the principles of the Declaration of Helsinki. All participants were informed about the study and asked to provide their names on a consent form. The study was registered with the University Hospital Medical Information Network (UMIN000050872).

## Clinical Trial

The clinical trial described in this paper was registered with the University Hospital Medical Information Network under the registration number UMIN000050872.

## Data availability statement

All data from this study are presented in this manuscript or are available from the corresponding author upon reasonable request.

## CRediT author statement

The author contributions are stated as follows: Y.M., G.T., and Y.K. drew the manuscript. Y.M., G.T., and Y.K. prepared the images. Y.M., K.S., Y.I., K.S., and Y.K. collected the patient information. K.N. and G.T were conducted statical analysis. Y.S., K.S., M.S., and Y.K. critically revised the manuscript to ensure intellectual content and supervision. All authors have approved all aspects of our work and have read and approved the manuscript.

## List of Supplemental Content

Online supplemental figures.docx

