## Supplemental Figure for "Benefits of hydrogen during constant load testing in healthy adults: A pilot double-blind randomized crossover trial"

This supplemental material has been provided by the authors to give readers additional information about their work.

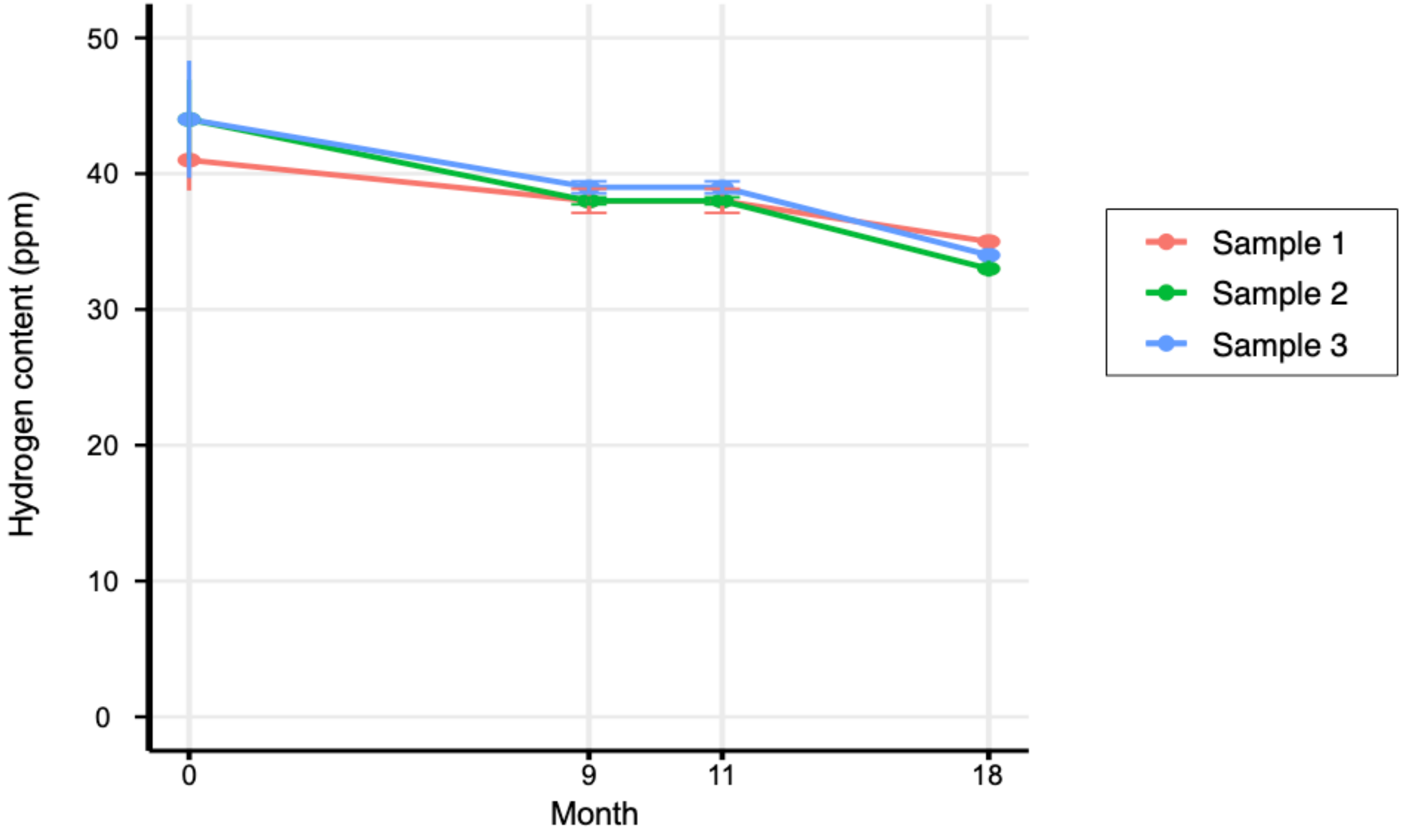


**Supplemental Figure 1. Hydrogen molecule content in hydrogen-rich jelly**

The concentration of dissolved hydrogen was measured using the headspace gas chromatography method and was confirmed to remain above 30 ppm for up to 18 months after production.

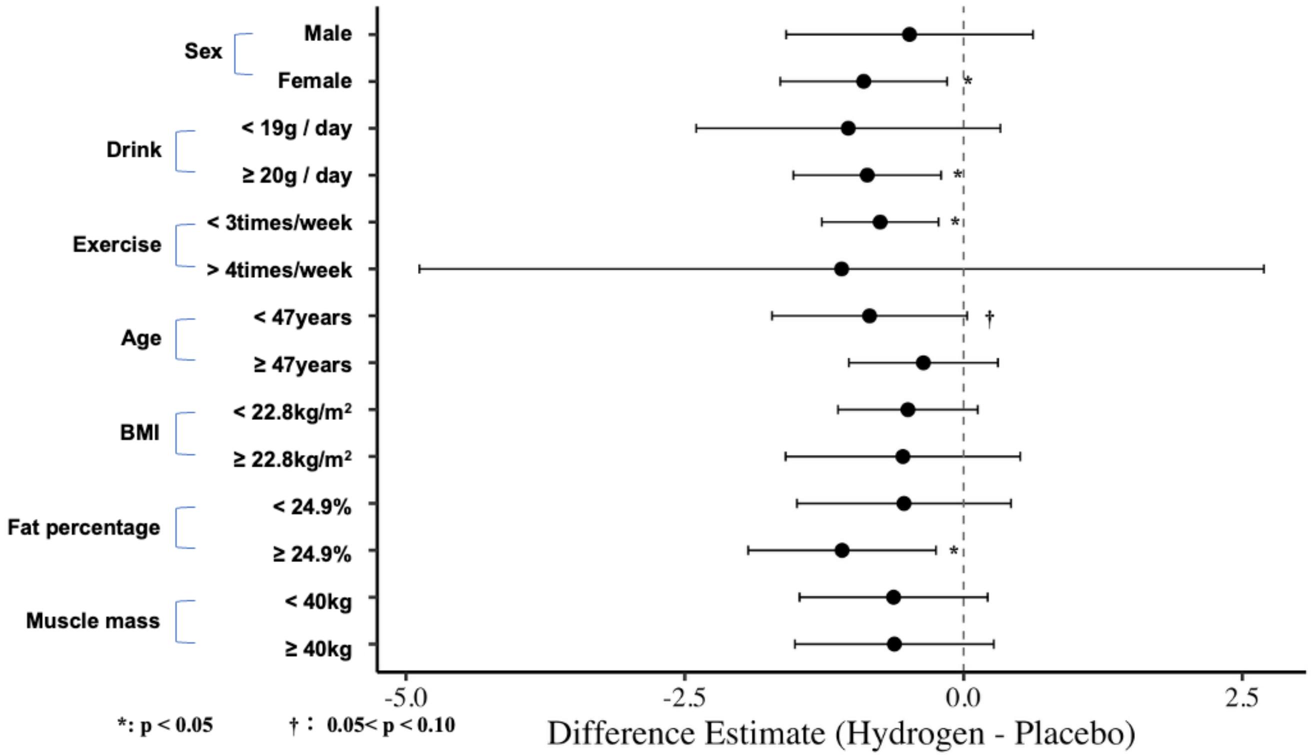


**Supplemental Fig. 2. Characteristics of participants who exhibited a reduction in peak blood lactate under the hydrogen-rich jelly (HRJ) condition.** Participants with lower peak blood lactate levels in the HRJ condition were characterized by female sex (mean difference = –0.90, 95% CI: –1.64 to –0.15, p = 0.023), daily alcohol consumption ≥20 g (mean difference = –0.86, 95% CI: –1.53 to –0.20, p = 0.014), exercise frequency less than three times per week (mean difference = –0.75, 95% CI: –1.27 to –0.23, p = 0.008), and body fat percentage ≥24.9% (mean difference = –1.09, 95% CI: –1.93 to –0.25, p = 0.018). In addition, participants under 47 years of age showed a trend toward greater reduction (mean difference = –0.84, 95% CI: –1.72 to 0.03, p = 0.058), although this was not statistically significant.

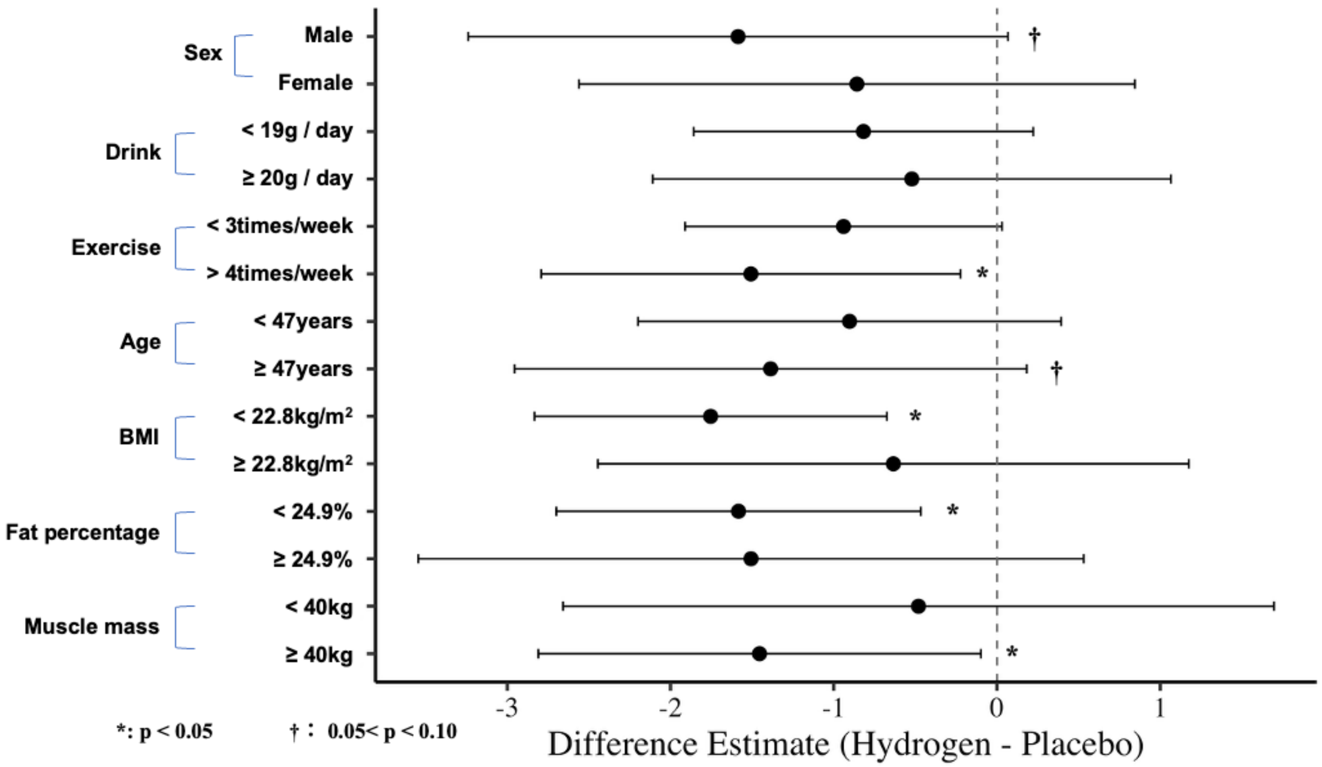


**Supplemental Fig. 3. Characteristics of participants who exhibited a reduction in peak oxygen uptake ($VO_2$) under the hydrogen-rich jelly (HRJ) condition.**

Participants with lower peak $VO_2$ in the HRJ condition were characterized by a body mass index (BMI) <22.8 kg/m² (mean difference = –1.75, 95% CI: –2.83 to –0.68, p = 0.004), body fat percentage <24.9% (mean difference = –1.58, 95% CI: –2.70 to –0.47, p = 0.009), skeletal muscle mass ≥40 kg (mean difference = –1.45, 95% CI: –2.81 to –0.10, p = 0.037), and habitual exercise frequency ≥4 times per week (mean difference = –1.51, 95% CI: –2.79 to –0.22, p = 0.033). In addition, male participants (mean difference = –1.59, 95% CI: –3.24 to 0.07, p = 0.059) and those aged ≥47 years (mean difference = –1.39,

95% CI: –2.96 to 0.18, $p = 0.075$) showed a trend toward greater reduction, although these did not reach statistical significance.
